# Genetic Metrics Decodes *Plasmodium falciparum* Diversity: Complexity of Infections, Parasite Connectivity, and Transmission Intensity in Mainland Tanzania’s Diverse Regions

**DOI:** 10.1101/2025.04.18.25326063

**Authors:** Dativa Pereus, Abebe A. Fola, Catherine Bakari, Misago D. Seth, Rebecca DeFeo, Beatus M Lyimo, Celine I. Mandara, Rashid A. Madebe, Doris D. Mbata, Zachary R. Popkin-Hall, Ramadhan Moshi, Ruth B. Mbwambo, Daniel Mbwambo, Sijenunu Aaron, Abdallah Lusasi, Samwel Lazaro, David J. Giesbrecht, Benard Kulohoma, Jonathan J. Juliano, Jeffrey A. Bailey, Gerald Juma, Victor A. Mobegi, Deus S. Ishengoma

## Abstract

**Background:** Recent initiatives have promoted the application of genomic data to determine the trends and patterns of malaria transmission and the impact of interventions. This study aimed to evaluate and identify the most effective genetic metrics for monitoring the genetic diversity of *Plasmodium falciparum* and its correlation with malaria transmission intensities in Mainland Tanzania.

**Methods:** A cross-sectional survey was conducted between February and July 2021 in 100 health facilities from 10 regions of Mainland Tanzania, categorized into four malaria transmission strata: high (two regions), moderate (two regions), low (three regions), and very low transmission strata (three regions). Dried blood spots (DBS) samples were collected from 12,875 symptomatic patients and all samples with positive results obtained by rapid diagnostic tests (n = 7,199) were sequenced using molecular inversion probes (MIPs). The MIPs targeted 1,832 single nucleotide polymorphisms (SNPs) distributed across the 14 *P. falciparum* chromosomes. Raw sequence data were analyzed using MIPTools and the filtered dataset was used to estimate different genetic metrics.

**Results:** After processing, 3,149 (43.0%) samples passed filtering conditions and were used for downstream analysis. The countrywide mean complexity of infection (COI) was 1.5, with 1,878 (59.6%) samples being monogenomic. The mean COI was significantly higher in high and moderate transmission strata (p < 0.001) compared to low and very low transmission strata. The odds of polyclonal infections were significantly lower in moderate (aOR = 0.67; 95% CI: 0.55-0.81; p < 0.001), low (aOR = 0.52; 95% CI: 0.43-0.63; p < 0.001), and in very low strata (aOR = 0.49; 95% CI: 0.40-0.61; p < 0.001) compared to the high transmission stratum. There was very little parasite genetic differentiation among regions with fixation index (*F_ST_*) values ranging from 0 to 0.006. The countrywide mean pairwise identity by descent (IBD) was 0.02. The mean pairwise IBD by transmission intensity was similar with mean IBD = 0.0155 in very low, 0.0158 in low, 0.0156 in moderate and 0.0152 in high malaria transmission strata. Using discriminant analysis of the principal component (DAPC) parasite populations from different regions clustered within regions suggesting genetic similarity among them.

**Conclusion:** Parasites from the sampled 10 regions had high complexity of infection and polyclonality, with a high correlation with regional malaria *transmission intensities*. Thus, these metrics can be potentially integrated with the current malaria surveillance and may be useful in the assessment of trends and patterns of malaria transmission. Further validation is needed to link these measures to current control strategies and evaluate how they might be used to determine the impact of different malaria transmission interventions in Mainland Tanzania.

## Introduction

Genomics surveillance, which combines epidemiological applications with genomics and bioinformatics methods, is becoming more recognized as a powerful tool for the surveillance of infectious diseases, including malaria (1,2). Malaria molecular surveillance (MMS), which has been endorsed by the World Health Organization (WHO) (3), is now being deployed in various endemic countries to complement traditional malaria surveillance efforts (2,4). MMS offers advantages over traditional methods by resolving local and regional transmission network; measuring the impact of interventions on parasite genomic diversity; and identifying sources and sinks of infections (1,5–8). It is also useful to detect the signatures of adaptation and natural selection (5,9); identify imported cases (5,8) and identify loci associated with vaccine escape (9,10), antimalarial drugs (11–13) or diagnostic resistance (14). Additionaly, MMS can be leveraged to track the spread of different parasite loci (11,15,16). Thus, MMS in combination with epidemiological data is increasingly becoming an essential tool for assessing programme performance, the impact of current interventions, and determining if there is a need for intervention adjustments as well as resource allocation for the targeted population(1,2,17).

Despite intensified interventions over the past two decades, the burden of malaria, mainly caused by *Plasmodium falciparum,* remains high in Sub-Saharan Africa (18). In 2023, Tanzania ranked fourth in malaria deaths, accounting for 4.3% of global malaria fatalities (19). Recent interventions have created varied transmission patterns in Mainland Tanzania, with some areas experiencing persistently high malaria burden while other parts of the country (40%) experience low to very low transmission intensities and disease burden (11,20,21). The country is stratified into four strata of different transmission intensities based on the prevalence of malaria in school children aged 5-16 years: very low (<1%), low (1 - <5%), moderate (5 - <30%), and high (≥30%) (20,22) (Figure 1A). Based on the WHO’s high-burden, high-impact initiative, the Tanzania National Malaria Control Programme (NMCP) targets these areas with tailored interventions to reduce the burden in high and moderate transmission areas and pursue elimination in regions with low and very low transmission intensities (21).

**Figure 1:**
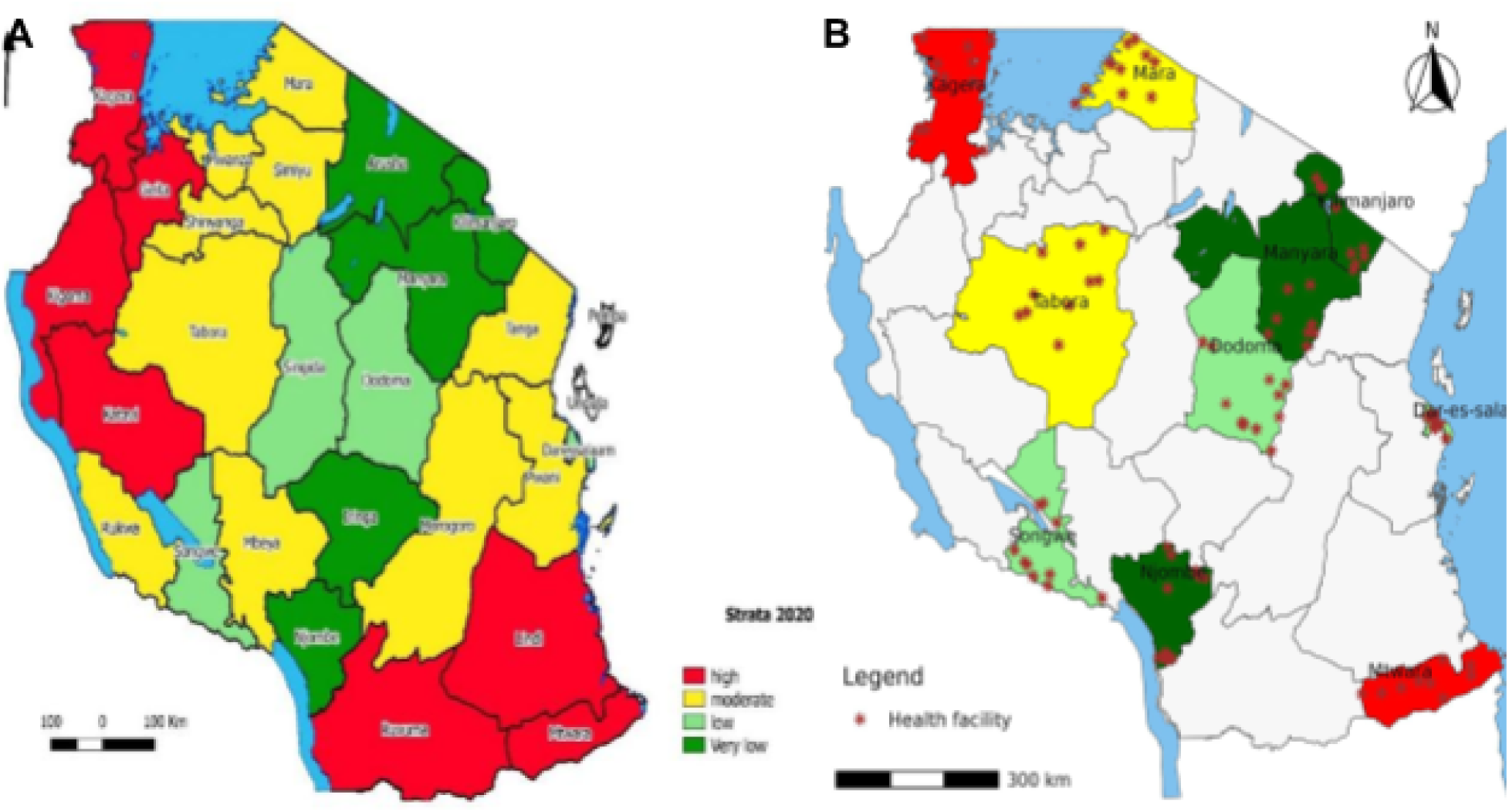
Maps of Tanzania showing the different strata of malaria transmission **(A)** and the MSMT study regions as well as sample collection sites **(B)**. In **(A)**, red show regions in high transmission stratum with prevalence in school children aged 5 - 16 years of >30%, yellow is for regions with moderate transmission with prevalence of 5-<30%, light green is for low transmission (prevalence of 1 - <5%) and dark green represent regions with very low transmission stratum (prevalence <1%) (Source: NMCP) (20). The red dots in Figure 1B show the health facilities which were sampled by the MSMT project in 2021 (12,30,33).

Until recently, most studies in Mainland Tanzania have generally analyzed small datasets and focused primarily on high-transmission regions (11). This limited the geographic scope and they could not provide a full exploration of the parasite genomic variations in many parts of Mainland Tanzania with varying transmission intensities. Studies in Tanzania are starting to focus on generating information about the patterns of transmission and disease burden to support the ongoing initiatives to appropriately modify intervention strategies, inform local control programmes, and apply targeted interventions. Leveraging both high-throughput genomic and epidemiological data, these studies help to provide baseline information on the patterns of parasite diversity, population structure, and drug resistance in high transmission regions on the mainland (5,11). Other studies have been conducted in Zanzibar, which has low transmission intensities and is currently implementing interventions for malaria elimination (5,7,8). Those studies reported varying patterns of parasite diversity (5,11,23) and signatures of selection (5,24) in both Mainland Tanzania and Zanzibar and provided evidence of the importation of malaria to Zanzibar from Mainland Tanzania (7,8). To date, these studies have not assessed how different genetic metrics can be leveraged to understand local malaria transmission or to to identify if these metrics can be utilized in routine surveillance systems for informed local control.

Recent studies undertaken in Tanzania and other endemic countries have leveraged Molecular Inversion Probes (MIPs), which is a high-throughput technique to capture, amplify, and sequence short-targeted regions of the *P. falciparum* genome that can be used to resolve different biological questions such as the parasite’s genetic diversity and transmission dynamics (11,25,26). MIPs are cost-effective and highly specific, as they use a targeted probe design. They can be performed directly from most sample types, including dried blood spots (DBS), thus making them implementable in routine malaria surveillance systems (11,25,26). MIPs have been successfully employed to study the spread of antimalarial drug resistance strains and parasite diversity and population structure in the Democratic Republic of Congo (26), Zambia (27), Ethiopia (28), in Mainland Tanzania (11–13) and Zanzibar (5,7,8). Thus, MIPs’ use in large-scale country-wide surveys, such as those undertaken by the project on molecular surveillance of malaria in Tanzania (MSMT) (15), can potentially and effectively resolve genomic variations of the *P. falciparum* populations circulating in different transmission strata within Mainland Tanzania.

This study aimed to evaluate and identify the most effective genetic metrics for monitoring the genetic diversity of *P. falciparum* and its correlation with changes in the transmission intensities in Mainland Tanzania. This study provides a high-resolution of parasite genetic diversity, which is critical for a better understanding of the contemporary parasite populations and transmission intensity across Mainland Tanzania. It supports NMCP policy and decision-making as Tanzania progresses towards malaria elimination by 2030.

## Methods

### Study sites and study design

The samples and metadata used in this study were collected from February to July 2021 through a country-wide cross-sectional survey which was conducted by the MSMT project. Details of study sites and sample collection procedures have been previously reported (14,15,29). Briefly, a cross-sectional survey of symptomatic patients was conducted at 100 health facilities (HFs) in 10 regions of Mainland Tanzania (Dar es Salaam, Dodoma, Kagera, Kilimanjaro, Manyara, Mara, Mtwara, Njombe, Songwe and Tabora) (14,30) (Figure 1B). According to the 2020 malaria stratification by NMCP, all 26 regions in Mainland Tanzania were grouped into four malaria transmission strata based on malaria prevalence in school children (aged 5 - 16 years), and the 10 regions sampled were drawn from all four strata (20) (Figure 1A).Two regions were located in high transmission (Kagera and Mtwara), two in moderate transmission (Mara and Tabora), three in low transmission (Dar es Salaam, Dodoma and Songwe) and three regions in the very low malaria transmission stratum (Kilimanjaro, Manyara and Njombe).

### Sample collection, DNA isolation and MIP sequencing

Procedures for sample collection at the HFs were described in previous MSMT publications (14,29,30). Briefly, febrile patients aged ≥6 months old with fever or a history of fever were tested for malaria using RDTs. A total of 12,875 patients were screened for malaria using rapid diagnostic tests (RDTs)(either Abbott Bioline Malaria Ag Pf/Pan (Abbott Diagnostics Korea Inc., Korea) or Smart Malaria Pf/Pan Ag Rapid Test (Zhejiang Orient Gene Biotech Co. Ltd, China) and 7,199 had positive results by RDTs. All RDT-positive samples were sequenced using MIPs and the data generated was further analyzed in this study (**Supplementaly Figure 1**).

DNA was extracted using the Chelex 100 + Tween20 protocol (15,33,34). MIP capture, sequencing, and PCR amplification were performed as described earlier (11,15,25–27) and targeted 1,832 SNPs in the 14 chromosomes of the *P. falciparum* genome. Gel electrophoresis was used to confirm capture success, and DNA libraries were pooled and purified using magnetic beads, followed by gel extraction (NEB). Quality control was performed using a Qubit™ Fluorometer and fragment analyzer. Libraries were denatured, diluted, and spiked with PhiX control before sequencing on the Illumina NextSeq 550 platform.

### Data management and bioinformatics analysis

Epidemiological and demographic data from study participants were collected at the health facility using paper questionnaires. After the survey was completed, the data were double-entered into Microsoft Access 2019 (Version 2302). Following data entry, the dataset was cleaned by removing duplicates, correcting mismatched information, eliminating samples with missing variables, and creating new variables using the provided dictionaries, as outlined previously (12, 34). The cleaned data were then merged with sequence data using “dplyr” package in R version 4.4.0 (The R Foundation for Statistical Computing, Vienna, Austria) for subsequent analysis, including descriptive statistics and estimation of genetic metrics within regions and across transmission strata. MIPtools and MIP Wrangler software were used to demultiplex and process the sequencing data, respectively. The processed data was then mapped to the P. falciparum 3D7 V3 reference genome using BWA (35,36).. Variant calling was performed using Freebayes, and the resulting vcf files were used for downstream analysis. Quality control was performed using bcftools software version 1.13, and only biallelic sites were retained (**Figure 2**).

**Figure 2:**
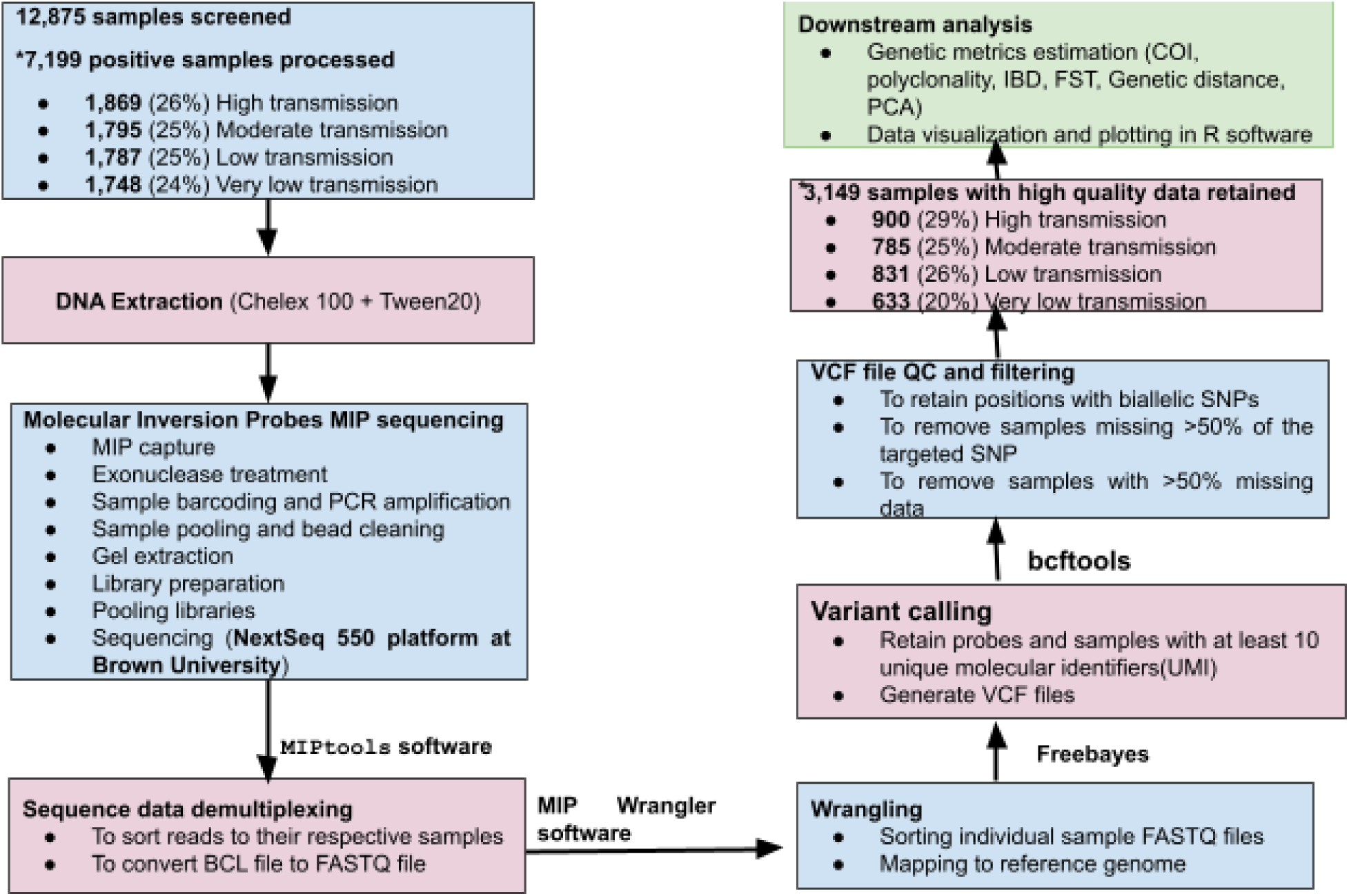
Flowchart showing how the data were generated and processed, and the downstream analysis performed. *\*The proportions of samples from regions located in the different strata of malaria transmission, which were sequenced (p=0.07). and those which passed the filtering criteria and were* used *in the analysis (p = 0.14) were not significantly different*.

### Estimation of within-host genetic diversity

Within-host parasite genetic diversity was measured by calculating the complexity of infection (COI) and the proportion of polyclonal infections. The COI is defined as the number of genetically distinct parasite strains in a single infection (37,38). The COI for each sample was calculated using a Markov Chain Monte Carlo method for estimating COI likelihood (*THE REALMcCOIL*) (39). The proportion of polyclonal infections was defined as the number of samples with COI > 1 over the total number of samples with successful genotypes, which was assessed in each region and transmission strata. Polyclonality was also assessed by age strate categorized into three groups; under five (<5 years), school children (5-<15 years) and adults (≥15 years).

The proportions of samples with polyclonal infections in the regions sampled were compared to the mean annual test positivity rates of each region using the same year’s data, which was obtained from the District Health Information System 2 (DHIS2) of the Ministry of Health. The “ggplot2” package was used to plot and visualize the distribution of COI per region, transmission strata and age group in R version 4.4.0.

### Analysis of parasite genetic relatedness and connectivity

The inbreeding_mle function of MIPAnalyzer (https://github.com/mrc-ide/MIPanalyzer) was used to detect relatedness across parasite populations by measuring the proportion of shared genome segments. IBD analysis measured relatedness among parasites by sharing ≥50% of the genome segments (half-siblings) or ≥90% (full-siblings/highly related) of their genome segments within and among the regions. The proportion of parasites’ pairwise IBD sharing was calculated for all samples and each region per transmission strata. Pearson’s chi-square test was used to test for significant differences in IBD sharing among regions and transmission strata.

### Parasite population structure and genetic differentiation

Evidence of parasite genetic differentiation among the study regions was assessed by calculating the pairwise Wright’s fixation index (*F_ST_*) using the “hierfstat” version 0.5-11 package (https://github.com/jgx65/hierfstat) and genetic distance using the ‘adegenet’ (V.2.1.10) package (https://github.com/thibautjombart/adegenet) in R version 4.4.0. To determine the population structure within and between *P. falciparum* populations in the regions (stratified into different strata of varying transmission intensities), Discriminant Analysis of Principal Component (DAPC) was performed using the “adegenet” (V.2.1.10) R package. The ggplot2 R package (v.3.4.0) was used to visualize the resulting DAPC plots.

### Association of the different genetic metrics with malaria transmission intensities

This analysis investigated the relationship between COI, polyclonality, IBD sharing and malaria transmission. Kruskal-Wallis and linear regression were used to assess the association of COI with transmission and age. Logistic regression was used to determine the association between polyclonal infections and transmission, and Pearson’s Chi-squared test was used to assess the association between IBD sharing and transmission. The results were considered significant when p<0.05 at 95% confident interval.

## Results

### Dataset and site information

We screened a total of 12,875 patients from 100 health facilities in 10 regions using RDTs and 7,199 (55.9%) were malaria positive by RDTs. All RDT-positive samples (n=7,199) were sequenced using MIP and after performing quality checks and filtering, 43.7% (n = 3,149/7,199) of the samples had good quality data and 55.2% (n = 1,011/1,832) of all the targeted SNPs had good quality (Supplementary table1). These samples were considered to have been successfully sequenced and were retained for downstream analysis. The proportions of samples with genotyping success (n = 3,149) from the four malaria transmission strata were not significantly different (p = 0.14), with 20.1% (n = 633/3,149) from regions located in the stratum with very low, 26.4% (n = 831/3,149) from low, 24.9% (n = 785/3,149) from moderate, and 28.6% (n = 900/3,149) from regions located in the high transmission stratum. Majority of the samples with high quality data were from the priority regions of Kagera (18.3%, n= 576/3,149), Tabora (13.0%, n = 410/3,149) and Mara (10.6%, n = 334/3,149) while the region with fewer samples was Kilimanjaro (3.6%, n= 114/3,149) (**Supplementary Figure 1 and Supplementary Table 1**).

### Variations of complexity of infections by transmission intensity and other variables

Within-host genetic diversity based on COI showed that over half of the samples (59.6%, n = 1,878/3,149) carried monoclonal infections (COI = 1), and 40.4% (n = 1,271/3,149) had polyclonal infections (COI > 1). The country-wide mean COI was 1.5, with the highest mean COI in Kagera (COI = 2.1, range = 1 - 22) and the lowest mean COI in Manyara (COI = 1.3, range = 1 - 4) (**Figure 3A**). Among the samples with polyclonal infections, 80.3% (n = 1,020/1,271) had COI = 2, 11.2% (n = 142/1,271) had COI = 3, and 8.3% (n = 105/1,271) had COI ≥4. Only a very small fraction of the samples (0.3%; n = 4/1,271) had COI ≥ 10. The mean COI was significantly higher in high (COI = 1.8, range = 1 - 22) and moderate (COI = 1.5, range = 1 - 6) transmission strata compared to low and very low transmission (p < 0.001), but it was similar in the regions within low (COI = 1.4, range 1 - 5) and very low transmission strata (COI = 1.4, range = 1 - 5) (p = 0.92) (**Figure 3B**). The mean COI was statistically different among regions and transmission strata (p < 0.001). Using the high transmission stratum as a reference group, the mean COI significantly decreased in other transmission strata, with the coefficient =-0.43 (95% CI:-0.53--0.33, p<0.001) in moderate,-0.52 (95% CI:-0.61--0.42, p<0.001) in low and the coefficient =-0.51 (95% CI:-0.61--0.41, p<0.001) in very low transmission strata.

**Figure 3.**
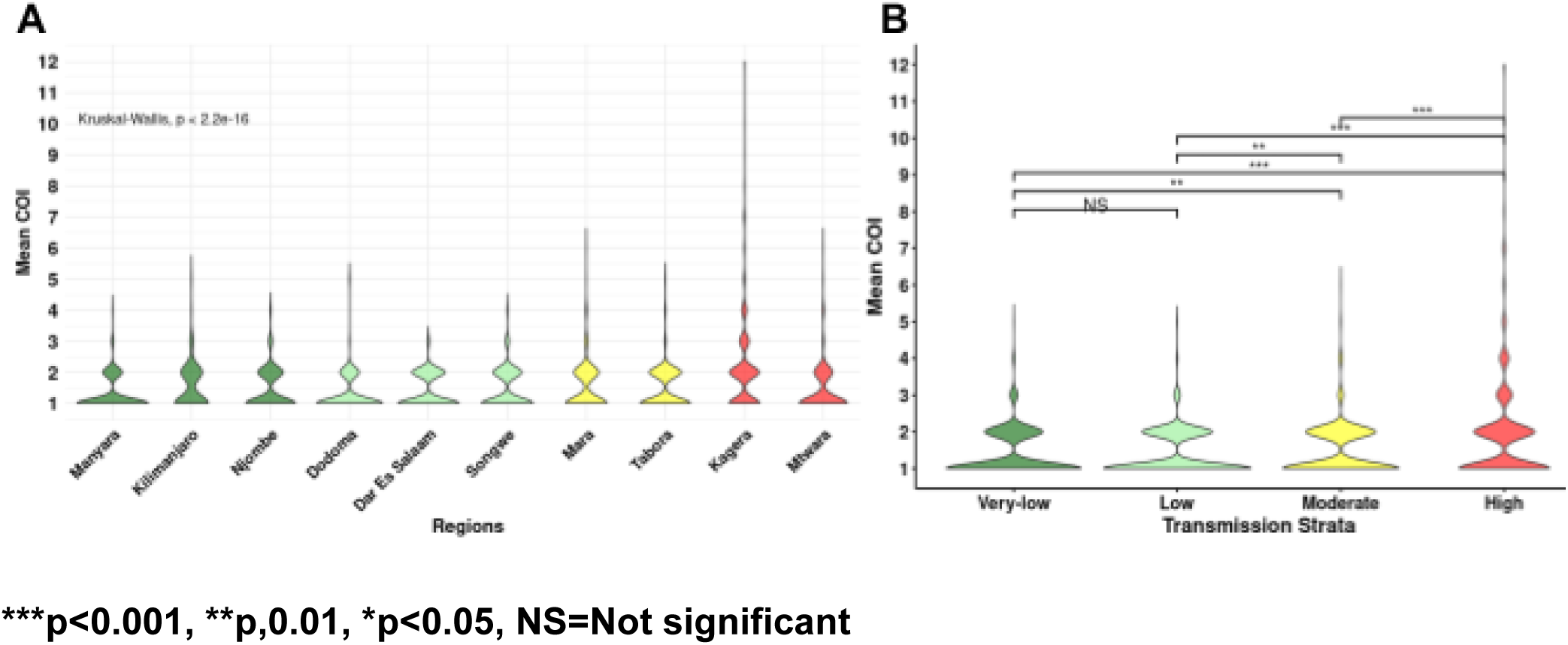
Violin plots show the distribution of the mean COI per region **(A)** and in different transmission strata **(B).** Dark green = regions in very low strata, green = regions in low strata, yellow = regions in moderate strata, and red = regions in high transmission strata.

The differences in mean COI among patients of different age groups were statistically significant (p < 0.001), with higher mean COI (1.6) among school children, followed by under-fives (COI = 1.5) and the lowest (COI =1.4) was in adult patients. The difference in mean COI was statistically significant between under-fives and adult patients (p = 0.003) and between school children and adult patients (p = 0.03). However, there was no significant difference in mean COI between under-fives and school children (p = 0.30). The mean COI was similar among female (COI = 1.6) and male (COI = 1.5) participants (p = 0.40) (**Supplementary Figure 2**).

### Spatial heterogeneity of polyclonal infections and transmission intensity

The countrywide proportion of polyclonality (COI > 1) was 40.4% (n = 1,271/3,149). Among all regions, the highest proportion of samples with polyclonal infections was in Kagera (61.3%, n = 353/576) and the lowest was in Manyara (23.4%, n = 52/222) as presented in **Figure 4A**. The proportion of samples with polyclonal infection in each region was relatively similar to the regional malaria test positivity rates based on 2021 data, except for Kilimanjaro which has fewer sequenced samples (**Figure 4B**). The proportion of samples with polyclonal infections was significantly higher in the high (50.4%; n = 454/900) and moderate transmission strata (40.4%; n = 317/785), compared to low (34.7%; n = 288/831) and very low strata (33.5%; n = 212/633, p < 0.001) (**Figure 5A**). Using the high transmission as a reference, the odds of polyclonal infections were significantly lower in moderate (aOR = 0.67; 95%CI: 0.55-0.81; p < 0.001), low (aOR = 0.52; 95%CI: 0.43-0.63; p < 0.001) and in very low transmission strata (aOR = 0.49; 95%CI: 0.40-0.61; p < 0.001). The proportion of polyclonal infections was 43.9% (n = 500/1,138) in under-fives (**Figure 5B**), 41.0% (n = 327/798) in school children (**Figure 5C**) and 36.6% (n = 444/1,213) among adult patients (**Figure 5D**). The odds of polyclonal infection were lower in adult patients compared to school children (aOR = 1.20; 95% CI: 1.00-1.45; p = 0.048) and under-fives (aOR = 1.36; 95% CI: 1.15-1.60; p < 0.001). The odds of polyclonal infections were not significantly different among males and females (aOR = 0.89; 95% CI: 0.77-1.03; p = 0.11) (**Supplementary Figure 3**)

**Figure 4:**
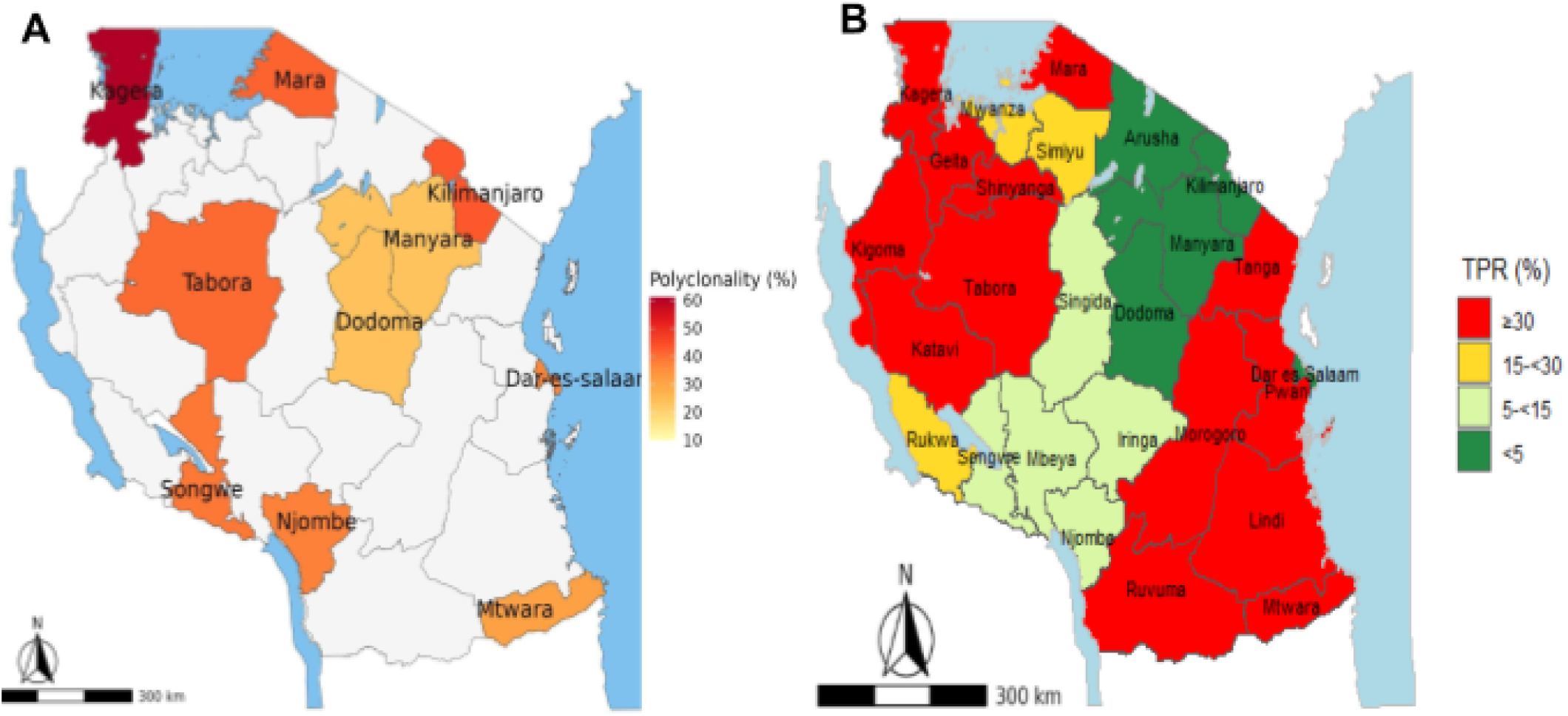
Maps of Tanzania showing the proportion of samples with polyclonal infections in the study regions **(A)** and the transmission intensity of malaria in Mainland Tanzania using the regional total positivity rate in 2021 **(B)**. The colours in Figure 5B are based on transmission intensities as described in Figure 1A, and the results showed that the proportions of polyclonal infections in the different regions were correlated with transmission intensities.

**Figure 5:**
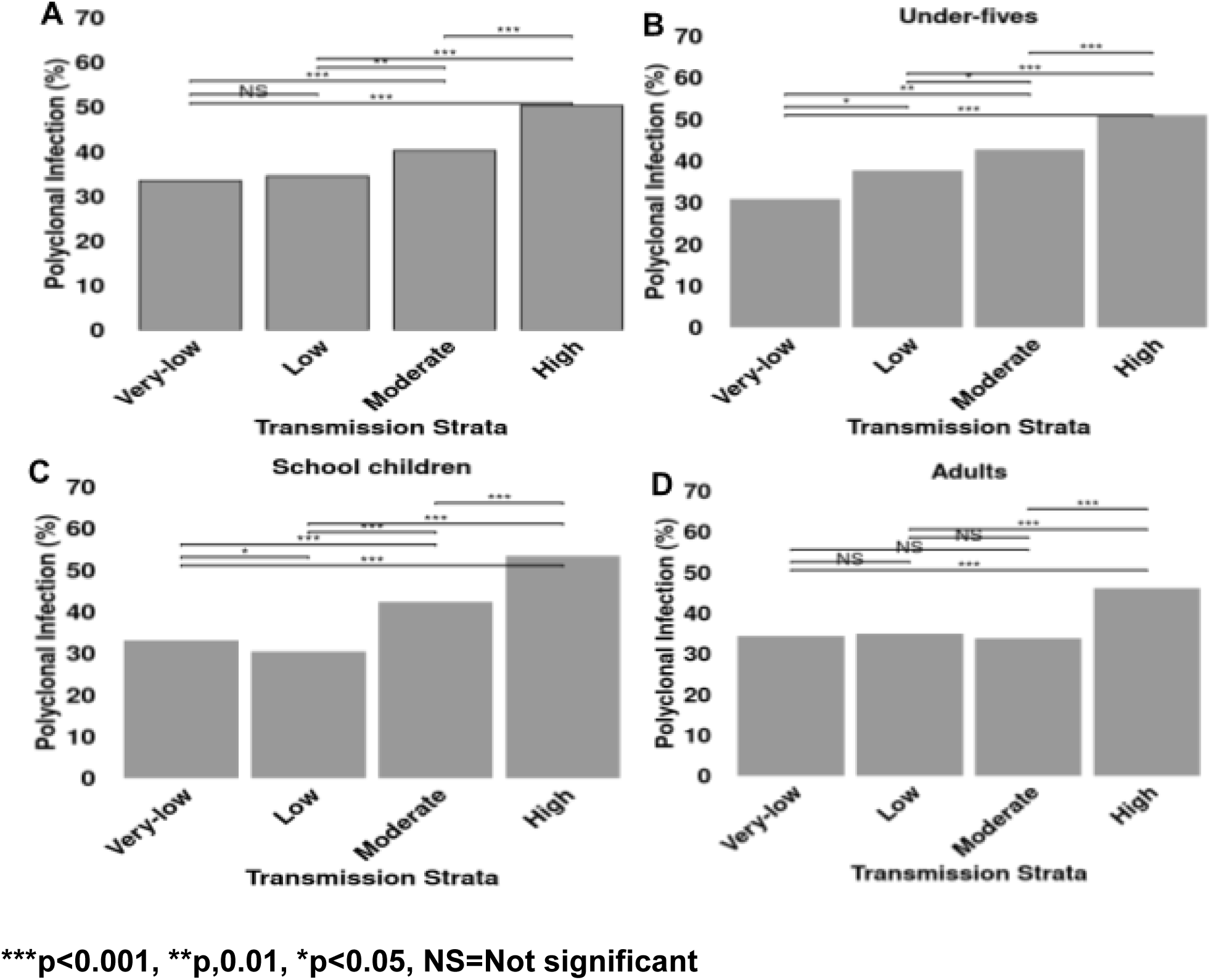
Histogram showing the proportion of polyclonal infections based on transmission strata **(A)** and different age groups (**B** = under-fives, **C** = School children and **D** = Adults)

### Parasite-relatedness and transmission intensity

The countrywide mean pairwise IBD was 0.02 among parasites populations. The mean pairwise IBD was similar across transmission strata (p = 0.39) with mean IBD = 0.0155 in very low, 0.0158 in low, 0.0156 in moderate and 0.0152 in high transmssion strata. There was no statistical difference in the mean IBD among regions (p = 0.44). The regional mean IBD ranged from 0.0148 in Kagera to 0.018 in Songwe region (Supplementary Table 2). Genetic relatedness among *P. falciparum* within regions was relatively high at ≥50% IBD sharing (half-siblings) (**Figure 6A**) and decreased at ≥90% IBD sharing (full-siblings/nearly clonal) (**Figure 6B**). In all 10 regions, the proportion of parasites sharing ≥50% IBD ranged from 7.6% in Tabora to 32.3% in Dodoma region and those sharing ≥90% IBD ranged form 2.75% in Tabora to 23.8% in Dodoma (**Figure 6C**).

**Figure 6:**
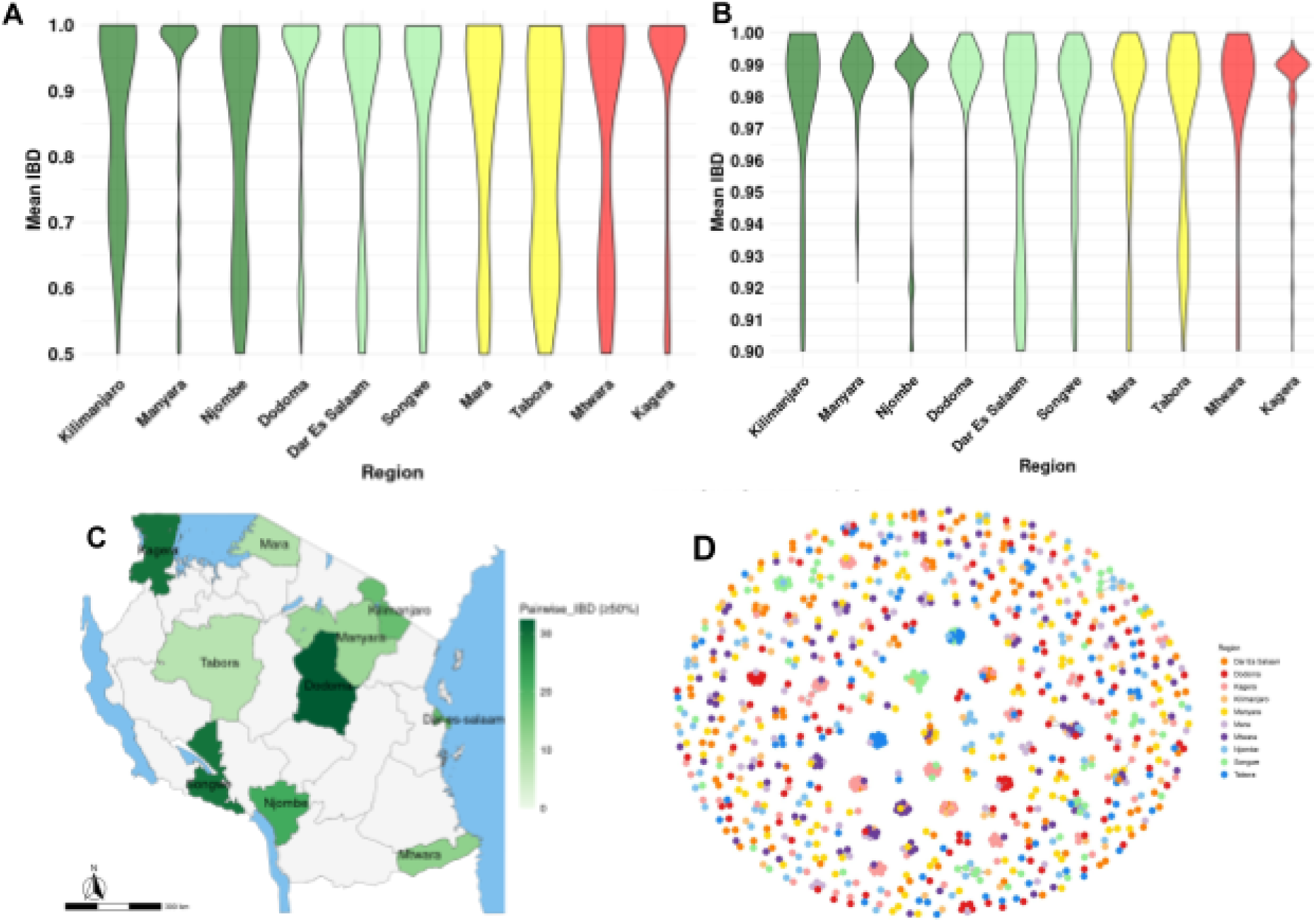
Violin plots showing the distribution of regional mean pairwise IBD for parasite sharing ≥50% (half-siblings) of their genome **(A)** ≥90% (full-siblings) of their genome per region **(B):** Darkgreen = regions in very low strata, green = region in low, yellow = regions in moderate and red regions in high transmission strata. (**C)** Map of Tanzania showing the proportion of parasite sharing ≥50% pairwise IBD in 10 regions. **(D)** IBD network showing parasites connectivity among regions.

The proportion of parasites sharing ≥50% pairwise IBD was 50.4% (n = 109/633) in the very low stratum, 79.8% (n = 233/872) in the low, 16% (n = 59/744) in the moderate, and 42.5% (n = 212/900) in high transmission strata. The proportion of parasites sharing ≥90% pairwise IBD was 30.2% (n = 64/633) in the very low stratum, 54.5% (n = 157/872) in the low, 7.5% (n = 27/744) in the moderate, and 29.2% (n = 149/900) in high transmission strata. The proportion of parasite sharing both ≥50% IBD and ≥90% IBD was significantly higher in low and very low transmission strata than in high and moderate (p<0.001). We also assessed if these parasites are connected across the country and found parasite populations among regions to be connected with parasites from Kilimanjaro and Manyara regions showing high connectivity with all other regions (**Figure 6D and supplementary Figure 4A**). We also found that IBD sharing is high among parasite within the same location and it decreases with increased geographical distances (**supplementary Figure 4B**)

### Parasite genetic differentiation and population structure

Based on the pairwise *F_ST_,* there was very little to no genetic differentiation among parasite populations in all regions, with *F_ST_* values ranging from 0 to 0.006 among the regions (**Supplementary** Figure 4). The *F_ST_*results showed that parasite populations from regions with a high geographical distance between them had higher *F_ST_* values compared to closer regions, suggesting that there was some level of genetic differentiation although the values did not reach the cut-off of *F_ST_* ≥0.05 which is significant.

Genetic distance analysis also revealed very little to no genetic distance among parasites across all regions with values ranging from 0 to 0.029 (**Figure 7A**). The DAPC analysis did not detect a clear, distinct parasite population clustering per region, except for parasites from the Kagera region that showed a low level of parasite clustering (**Figure 7B**). Samples within the same region clustered together, indicating within-region parasite genetic similarity. Different parasite populations formed overlapping clusters, indicating limited genetic diversity among parasites circulating within Mainland Tanzania

**Figure 7:**
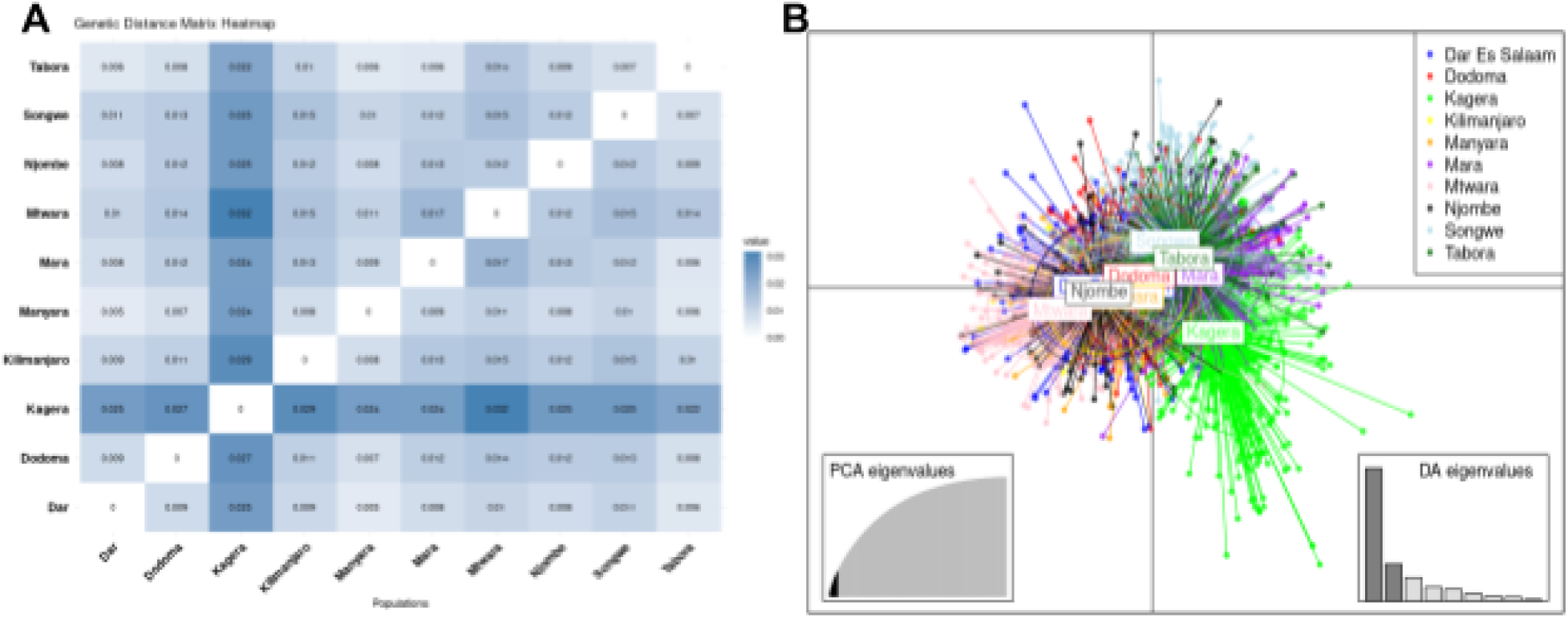
The heatmap shows parasite genetic distance values among regions using the genetic distance matrix **(A)**, and the DAPC plot shows the countrywide parasite population structure with colours representing parasites for each region **(B)**. The bar chart on the bottom left represents the eigenvalues, which show genetic variations explained by each PCA. The bar chart on the bottom right represents the contribution of the discriminant axes (DA) to the clustering, with the first two axes contributing most to the variation explained by the PCA.

## Discussion

This study aimed to evaluate various genetic metrics, analyze their correlation with malaria transmission intensity, and identify the most effective metric for monitoring *P. falciparum* genetic diversity and its potential use in malaria surveillance in Mainland Tanzania. This was the largest and first of its kind in Mainland Tanzania that covered all four malaria transmission strata. It used data and samples collected from 10 regions across four transmission strata, offering a comprehensive view of the parasite’s genetic landscape, based on the largest genomic dataset ever collected from the MSMT project.

The study applied a range of genetic metrics to examine genetic diversity, relatedness, and population structure in parasite populations in 10 regions. The findings provided deeper insights on the usefulness of genetic metrics (COI, proportion of polyclonality and IBD) in ressolving transmission intensity in areas with different levels of endemicity in Mainland Tanzania. The study used all samples with RDT-positive results without microscopy and qPCR data to perform sequencing using MIP, which enabled the generation of the data within a short time and provided important evidence to the Tanzanian NMCP as previously reported (12). This study adds important evidence on the genomic diversity of *P. falciparum* in Tanzania and the informativeness of different genetic metrics which can potentially be useful in malaria surveillance as part of the ongoing elimination efforts in Tanzania.

This study analysed a large number of samples (n = 7,199) with positive RDT results, although the success rate was lower and only 43.7% of the samples were retained for downstream analyses after filtering. Still, this is among the largest datasets for a country-wide study and the lower success rate was not unexpected. It has been extensively shown that RDTs which detect histidine-rich protein 2 (HRP2) have high rates of false positive results due to the persistence of the antigens after parasite clearance by drugs or the immune system (40). Previous studies also showed that some of the RDT-positive samples have low parasitaemia which may be difficult to sequence (41), especially in low and very low transmission settings. In addition, MIP sequencing utilizes probes which are applied to raw samples before amplification, making this approach have a low detection and success rate compared to the methods that work with amplified PCR products. The success rate is expected to be low in samples with varying levels of parasitaemia, such as those which were analysed in this study, since they were only RDT positive with unknown levels of parasitaemia. Despite these limitations, MIP sequencing proved to be a quick and relatively inexpensive method for analysing large volumes of samples and generating timely results as reported earlier (12).

The study revealed high within-host parasite diversity, indicated by elevated COI and a high proportion of polyclonal infections. Parasites showed high genetic relatedness and minimal differentiation based on *F_ST_*and genetic distance. As in previous studies (11,20,22), mean COI and polyclonality were strongly associated with transmission intensity with highest in regions with moderate to high malaria transmission, where frequent superinfection and co-transmission promote genetic diversity through recombination (1,4,42). In contrast, low and very low transmission areas had significantly lower COI and polyclonality, likely due to reduced exposure to diverse strains and limited transmission events. These trends align with earlier findings and highlight the potential of COI, polyclonality, and IBD as robust metrics for tracking malaria transmission dynamics and evaluating intervention impact in Mainland Tanzania and beyond. Notably, nearly half of the infections were polyclonal (40.4%), with rates reaching 50% in higher transmission regions. In low transmission settings, lower immunity may also lead to symptomatic cases being treated early, further reducing the opportunity for multiple strain infections (11,46).

Recent reports showed that some regions experience a high burden while others have persistently reported low rates of transmission intensities for about 20 years (20,22,47). Among the regions within the very low transmission strata, Kilimanjaro had a high mean COI of 1.52, and the proportion of polyclonal infections was 42.9%. This region had few samples that passed the filtering conditions. This indicated that most of the infections in this region were due to multiple parasite strains contributing to high diversity. However, in very low malaria settings, multiple parasite infection is not common and these findings may suggest that there is ongoing importation of genetically distinct parasite strains to these regions from areas with moderate and high transmission areas by travellers and human migrants. Looking at **Figures 6D** and **7B** parasites from Kilimanjaro were highly connected to parasites from other regions and in the PCA they are tightly clustered with other regions. Kilimanjaro is a tourist region and has a high movement of people. Importation in very low malaria settings has been reported to contribute to high genetic diversity in these settings as it was highlighted in different studies conducted in Zanzibar (5,7,8), Zambia (27) and the Kingdom of Eswatini (48).

There were significant differences in mean COI among age groups, with the highest mean COI among under-fives and school-aged children compared to adults across all transmission strata. High mean COI and *P. falciparum* genetic diversity among under-fives and school children has been reported in most endemic countries, including Zambia (49), Kenya (50), Tanzania (10) and Cameroon (51). A high rate of polyclonal infections among younger children could be influenced by variability in their level of acquired immunity (52). In malaria-endemic areas, immunity against malaria develops with age due to exposure to malaria infections that depend on the level of transmission intensities (53). Underdeveloped immunity can allow different strains of parasites to be detected in an infection, while fully developed immunity can help to clear certain strains leading to lower complexity of infections (54). In addition, recent studies from country-wide surveys have shown that the prevalence of malaria infections in most parts of Tanzania is higher among school children and under-fives in some areas (29,30,55–57), which could explain the high complexity of infections in these groups. Additionally, a country-wide analysis of subpatent infections has reported a higher prevalence of such infections in under-fives and school children, further supporting the observed high complexity of infections in these children (58). Thus, future studies will be needed to further explore the causes of the high complexity of infections in children and how this pattern responds to interventions currently directed to these groups.

Parasite relatedness, measured by pairwise IBD, varied across regions and transmission strata. The high-level parasite population connectivity and relatedness observed within regions, especially those from low and very low transmission strata, suggest a high rate of co-transmission of closely related parasite populations and a fingerprint of reduction in transmission intensities in some parts of the country. Similar findings have been reported in different studies that showed high parasite-relatedness in regions with low and very low transmission intensities, which is associated with the circulation of clonal parasites (4,6,7,27,43,59).

In areas with moderate to high transmission, increased recombination events, polyclonal infections, and rapid variations among *P. falciparum* population have been reported to contribute to reduced IBD sharing as the High-level parasite population connectivity and relatedness observed within regions, especially those from low and very low transmission strata, suggest a high rate of co-transmission of closely related parasite populations and a fingerprint of reduction in transmission intensities in some parts of the country an indication of high ongoing transmission in those areas (60,61). However, the proportion of parasite-relatedness within the Kagera region which is in a high transmission stratum was high suggesting a high rate of ongoing local transmission within the region. Despite observing a high mean COI in the Kagera a high rate of relatedness based on IBD sharing indicated that most infections within the region are due to co-transmission rather than superinfection that influences the closely related co-infecting strains within the population. However, parasites from Kagera showed relatively high *F_ST_* and genetic distance values (**Figure 7A and Supplementary** Figure 5) that may be influencing the observed high IBD sharing in this region. Parasite connectivity among *P. falciparum* populations was generally high across regions suggesting challenges for regional malaria elimination efforts in Mainland Tanzania. It was also shown that pairwise IBD sharing between parasites decayed with increasing geographical distances and there was high relatedness among parasites within the same geographical areas compared to between areas (supplementary figure 4), suggesting that IBD is a robust metric to capture local transmission dynamics and is not affected by allele frequency variability (8,59,62,63). This is possibly due to decreased inbreeding rates among *P. falciparum* populations between regions, thus adhering to the rule of isolation by distance. Decay in IBD sharing by geographical distances was previously reported in the Democratic Republic of the Congo (DRC) (26), Senegal (59), and Zanzibar (5,7,8), where IBD values were higher within clusters than between clusters. In addition, these findings suggest a low genetic diversity within the regions with low and very low transmission intensities, which is associated with higher inbreeding rates among the *P. falciparum* populations. ated with higher inbreeding rates among the P. falciparum populations.Notably, parasite populations from different regions clustered together in a DAPC, showing overlapping clusters indicating gene flow and shared genetic diversity among them. For example, parasites from Kilimanjaro, Dodoma, Manyara, Dar es Salaam, and Njombe were tightly clustered together (Figure 8). Also, parasite populations from Songwe, Mara, and Tabora overlapped. This indicates high parasite movements across Mainland Tanzania due to the recent upsurge of high mobility of human hosts and potentially vectors as well. The overlapping clusters observed in Mainland Tanzania indicate that there is a high level of gene flow and shared ancestry, and parasites are highly interconnected, accelerating the mixing of genetic information from different populations influencing parasite-relatedness. Also, human movement, trade and other economic activities from areas with different transmission intensities may be contributing to the introduction of different parasite strains in the population triggering the transmission of related parasite populations. However, parasite populations from very low transmission areas (Kilimanjaro, Manyara and Njombe) clustered with parasites from moderate and high transmission strata, a situation that may indicate parasite importation in these regions as importation in these settings have repeatedly been reported in different countries (5,7,8,27,48). However, more studies incorporating travel data will be needed to further understand the patterns and causes of parasite importation, relatedness, and connectivity, and how to use such data for malaria control and elimination in areas of low transmission intensities. Parasites from the Kagera regions formed a distinct cluster from the rest, indicating some levels of genetic differentiation compared to parasites from other regions.

Countrywide parasite genetic differentiation was very low and not significant. The estimated *F_ST_* and genetic distance values were very small, indicating very little to no genetic differentiation. However, *F_ST_* values showed that parasites from regions with very high geographical distances between them experienced slightly higher values compared to very close regions. Among the 10 regions, parasites from the Kagera region exhibited relatively high genetic distances when compared to other regions. These findings indicate that highly geographically isolated regions may have distinct populations, but the differences in *F_ST_*values were not significant, possibly due to high genetic diversity in these regions, which is linked to sustained high transmission intensities. Lack of genetic differentiation is also supported by the DAPC which showed overlapping clusters among the parasite population. These findings were similar to studies that have reported a lack of clear population structure based on DAPC in different transmission settings (25,26,44,59,64). This further supports the results from the FST and genetic distance matrix that show parasites to be related and lack genetic differentiation, suggesting clonal expansion of closely related parasites.

However, in a previous study that used MIPs data from a few regions (11), the DAPC plots showed clear clusters among parasites from different localities. This is different from the DAPC results of this study, which didn’t show clear clusters despite the data being collected from many regions with varying transmission intensities. The distinct cluster formed by parasites from Kagera may be due to limited gene exchange and geographical isolation that may trigger some parasite clustering alone, not mixing with parasite populations from other regions. However, the distinct cluster for parasites from Kagera may be linked to the recent evidence of partial artemisinin resistance in the region (12,13), which may be triggering the observed isolation by the parasite from this region that indicates genetic variability among parasite strains in the region. Further studies are warranted to investigate the genetic factors contributing to the clustering of parasites in this region, enhancing our understanding and guiding more effective intervention.

### Limitations of the study

This study used data from cross-section surveys to study different population genetics parameters of parasites circulating in different areas of Mainland Tanzania during that sampling time. These results cannot reveal any temporal nature or patterns of parasite populations in the country. Thus, longitudinal surveys with data collection covering different seasons are recommended in future studies. In addition, sampling techniques were not random because surveys were done in health facilities that had a sufficient number of cases and were capable of attaining the required sample size. Furthermore, this analysis focused on spatial scale and could not resolve the temporal resolution of the parasite genetic diversity in Mainland Tanzania. This will be done in the ongoing analysis to provide more insights into how parasite genetic diversity changes with time.

## Conclusion

This country-wide genomic analysis revealed high genetic diversity, spatial patterns of connectivity and limited population structure among *P. falciparum* populations in Mainland Tanzania. The mean COI and the proportion of polyclonal infection revealed a high correlation with transmission intensities and can be potentially used to assess trends and patterns of malaria transmission and evaluate the impact of malaria interventions in Mainland Tanzania. Further validation is needed to link these measures to specific interventions. They can be integrated into routine surveillance to resolve transmission dynamics and assess the effectiveness of interventions. Also, this study proves the beneficial use of molecular surveillance approach that covered all four transmission strata, providing a broader picture of the *P. falciparum* genetic diversity and current transmission intensity. As the country experiences high to very low transmission intensities, further research and analysis covering the entire country should be done to understand the genetic characteristics of circulating parasites and develop appropriate control interventions for each strata across the country.

## Ethical approval

The MSMT study protocol was approved by the Tanzania Medical Research Coordinating Committee (MRCC) of the National Institute for Medical Research (NIMR) and included approved standard procedures for informed consent and sample identification. Approvals to conduct this study in the respective regions were sought from the President’s Office, Regional Administration and Local Government (PO-RALG), and Regional Administration Secretaries (RAS) of each region oversaw the project at the regional level.

## Author’s contributions

DP and DSI formulated the original idea. DSI, CIM, RAM, DP and CB conducted the field surveys. DP, AAF, DJG, AS, BML, DR, MDS, RAM and CB performed data generation and computational analysis under the guidance of DSI, JJJ and JAB. DP wrote the first draft. DP, AAF, JJJ, AS, AF, RB, FF, RBM, RM, DM, VM, GJ, DM SA, AL, BK, SL, DJG, BML, ZPH, CB, JRG, MDS, CIM, JAB, and DSI reviewed and edited the manuscript. All authors contributed to the article and approved the submitted version.

## Supporting information

https://docs.google.com/document/u/0/d/1J8Dih9BMFV91-nBbYq75lFNUoK5nyP-aX4VpHkoIubw/edit

## Data Availability

All data are available and can be obtained from the project principal investigator upon a reasonable request

## Acknowledgements

We would like to express our gratitude to the participants and the parents or guardians of all children who participated in the surveillance. We acknowledge the contribution of the MSMT project staff and other colleagues who participated in the data and sample collection and laboratory processing of samples: Raymond Kitengeso, Ezekiel Malecela, Muhidin Kassim, Athanas Mhina, August Nyaki, Juma Tupa, Anangisye Malabeja, Emmanuel Kessy, George Gesase, Tumaini Kamna, Grace Kanyankole, Oswald Osca, Richard Makono, Ildephonce Mathias, Godbless Msaki, Rashid Mtumba, Gasper Lugela, Gineson Nkya, Daniel Challe, Richard Malisa, Sawaya Msangi, Ally Idrisa, Francis Chambo, Kusa Mchaina, Neema Barua, Christian Msokame, Rogers Msangi, Salome Simba, Hatibu Athumani, Mwanaidi Mtui, Rehema Mtibusa, Juma Akida and Tilaus Gustav. We are grateful to the finance, administrative and logistic support team at NIMR: Christopher Masaka, Millen Meena, Beatrice Mwampeta, Gracia Sanga, Neema Manumbu, Halfan Mwanga, Arison Ekoni, Twalipo Mponzi, Pendael Nasary, Denis Byakuzana, Alfred Sezary, Emmanuel Mnzava, John Samwel, Daud Mjema, Seth Nguhu, Thomas Semdoe, Sadiki Yusuph, Alex Mwakibinga, Rodrick Ulomi and Andrea Kimboi. We also wish to thank the management of the National Institute for Medical Research, the National Malaria Control Program and the President’s Office-Regional Administration and Local Government (including the Regional Administrative Secretaries of the 14 regions, district officials, staff from all 100 HFs and community health workers from the three community cross-sectional survey regions) for supporting the project. Financial and logistics support from the Bill and Melinda Gates Foundation team is highly appreciated.

## Data availability

All data and codes are available and can be obtained from the project principal investigator upon a reasonable request.

## Permission to publish

Permission to publish this manuscript was sought and obtained from the Director General of NIMR and the chairperson of the Tanzania Medical Research Coordinating Committee (MRCC).

Supplementary files

