## Supplementary material for "Genetic Metrics Decodes *Plasmodium falciparum* Diversity: Complexity of Infections, Parasite Connectivity, and Transmission Intensity in Mainland Tanzania’s Diverse Regions": https://docs.google.com/document/u/0/d/1J8Dih9BMFV91-nBbYq75lFNUoK5nyP-aX4VpHkoIubw/edit

**SUPPLEMENTARY FILES**

**Table 1. Summary of samples in each of the study regions stratified by transmission strata**

| Transmission strata | Region | Negative RDT(n) | Positive RDT(n) | Malaria prevalence (%) | Samples with good quality(%) |
|---|---|---|---|---|---|
| High | Kagera | 553 | 986 | 986/1,539 (64.1) | 576/986 (58.4) |
| | Mtwara | 297 | 883 | 883/1,180 (74.8) | 324/883 (36.7) |
| **Sub-total** | | **850** | **1,869** | **1,869/2,719 (68.7)** | **900/1,869 (48.2)** |
| Moderate | Tabora | 233 | 868 | 868/1,101 (78.8) | 410/868 (47.2) |
| | Mara | 437 | 927 | 927/1,364 (68.0) | 334/927 (36) |
| **Sub-total** | | **670** | **1,795** | **1,795/2,465 (72.8)** | **744/1,795 (41.4)** |
| Low | Dar-es-salaam | 241 | 572 | 572/813 (70.4) | 266/572 (46.5) |
| | Dodoma | 417 | 594 | 594/1,011 (58.8) | 235/594 (39.6) |
| | Songwe | 269 | 621 | 621/890 (69.8) | 371/621 (59.7) |
| **Sub-total** | | **927** | **1,787** | **1,787/2,714 (65.8)** | **872/1,787(48.8)** |
| Very-low | Kilimanjaro | 2654 | 615 | 615/3269 (18.8) | 114/615 (18.5) |
| | Manyara | 292 | 562 | 562/854 (65.8) | 222/562 (39.5) |
| | Njombe | 283 | 571 | 571/854 (66.9) | 297/571 (52) |
| **Sub-total** | | **3229** | **1,748** | **1,748/4,977 (35.1)** | **633/1,748 (36.2)** |
| **Total** | | **5,676** | **7,199** | **7,199/12,875 (55.9)** | **3,149/7,199(43.7)** |

**Table 2: Summary of regional mean IBD sharing and the proportion of parasite sharing ≥50% IBD and ≥90% IBD across transmission strata**

| Region_name | Mean_IBD | Total samples | IBD≥50 (n) | Proportion_IBD ≥50 (%) | IBD≥90 (n) | Proportion_IBD ≥90 (%) | Transmission _strata |
|---|---|---|---|---|---|---|---|
| Kagera | 0.0148 | 576 | 170 | 29.51 (170/576) | 124 | 21.53 (124/576) | High |
| Mtwara | 0.0166 | 324 | 42 | 12.96 (42/324) | 25 | 7.72 (25/324) | |
| Dar Es Salaam | 0.0149 | 266 | 48 | 18.05 (48/266) | 32 | 12.03 (32/266) | Low |

| | | | | | | | |
|---|---|---|---|---|---|---|---|
| Dodoma | 0.0159 | 235 | 76 | 32.34 (76/235) | 56 | 23.83 (56/235) | |
| Songwe | 0.0181 | 371 | 109 | 29.38 (109/371) | 69 | 18.60 (69/371) | |
| Mara | 0.0151 | 334 | 28 | 8.38 (28/334) | 16 | 4.79 (16/334) | Moderate |
| Tabora | 0.0162 | 410 | 31 | 7.56 (31/410) | 11 | 2.68 (11/410) | |
| Kilimanjaro | 0.0157 | 114 | 20 | 17.54 (20/114) | 12 | 10.53 (12/114) | Very low |
| Manyara | 0.0154 | 222 | 25 | 11.26 (25/222) | 19 | 8.56 (19/222) | |
| Njombe | 0.0153 | 297 | 64 | 21.55 (64/297) | 33 | 11.11 (33/297) | |

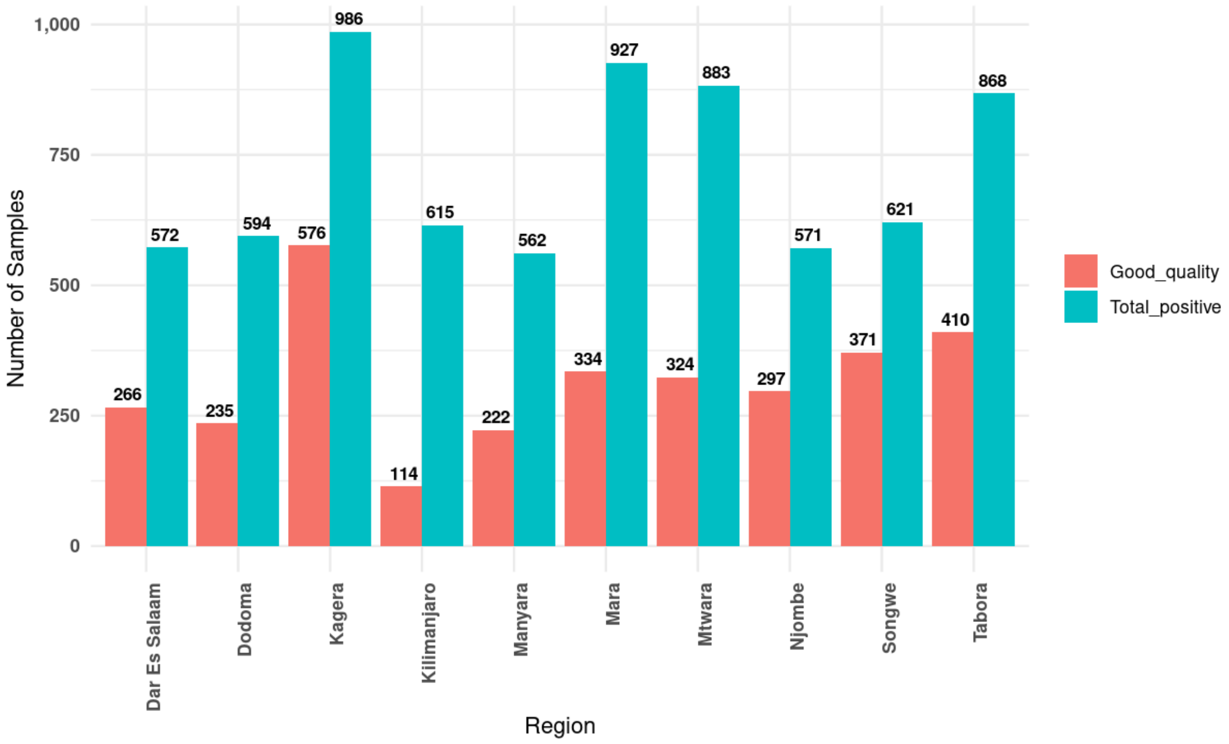


**Figure 1:** Histogram showing the regional distribution of samples sequenced(sky blue colour) and samples with good quality(salmon colour).

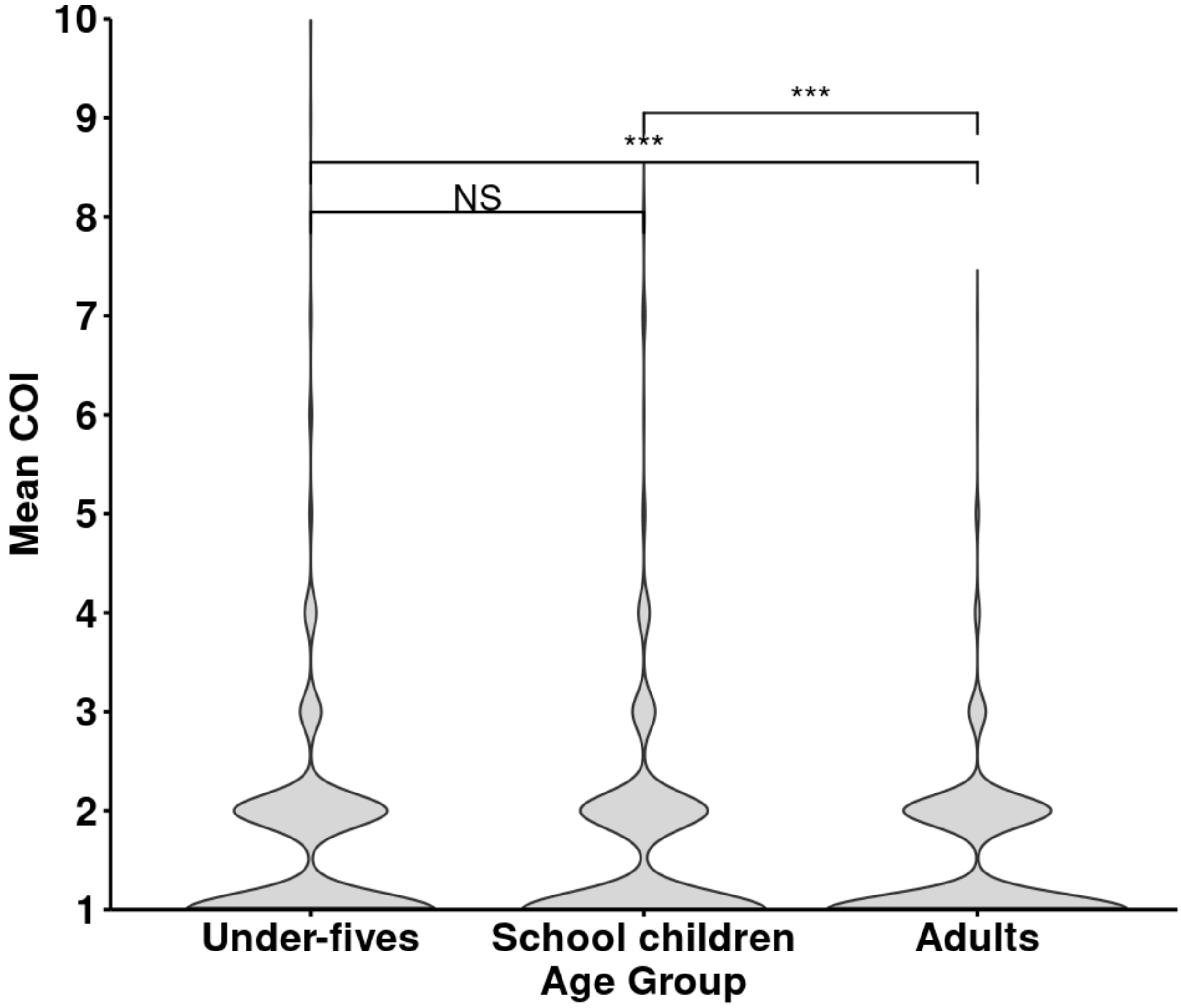


**Figure 2:** violin plots showing the distribution of mean COI per age group

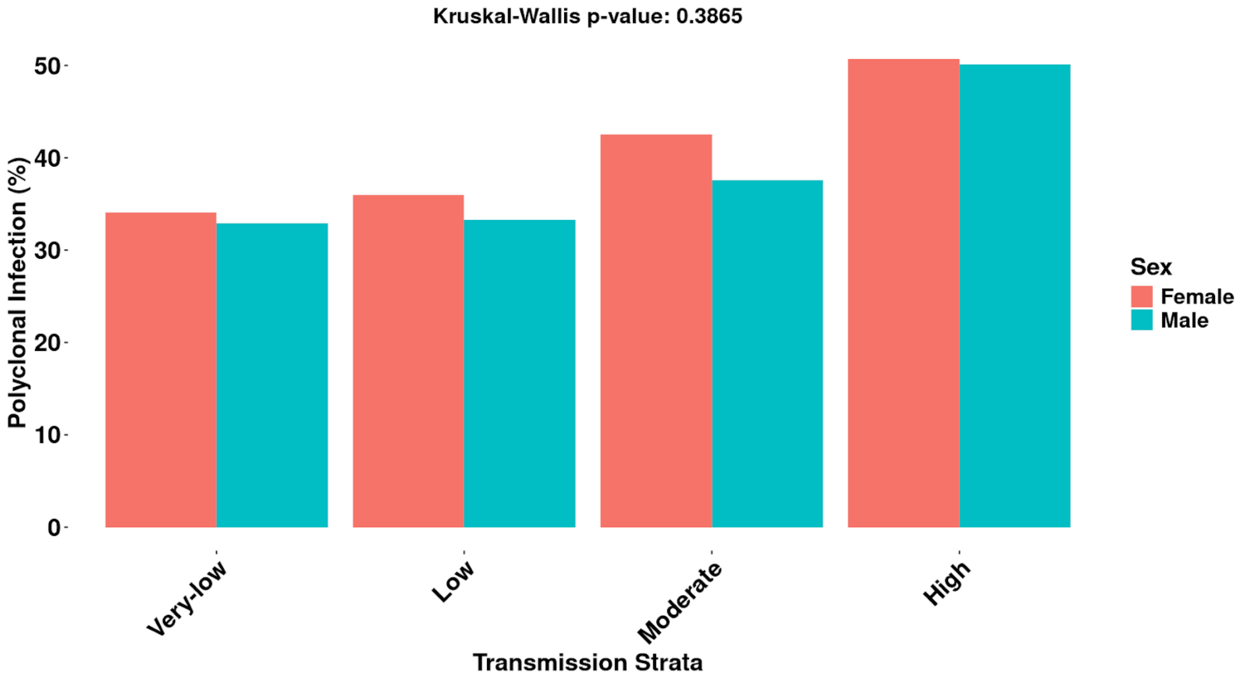


**Figure 3:** Bar plots showing the proportion of polyclonal infections by sex among transmission strata. Sky blue colour represents male and salmon colour represents females.

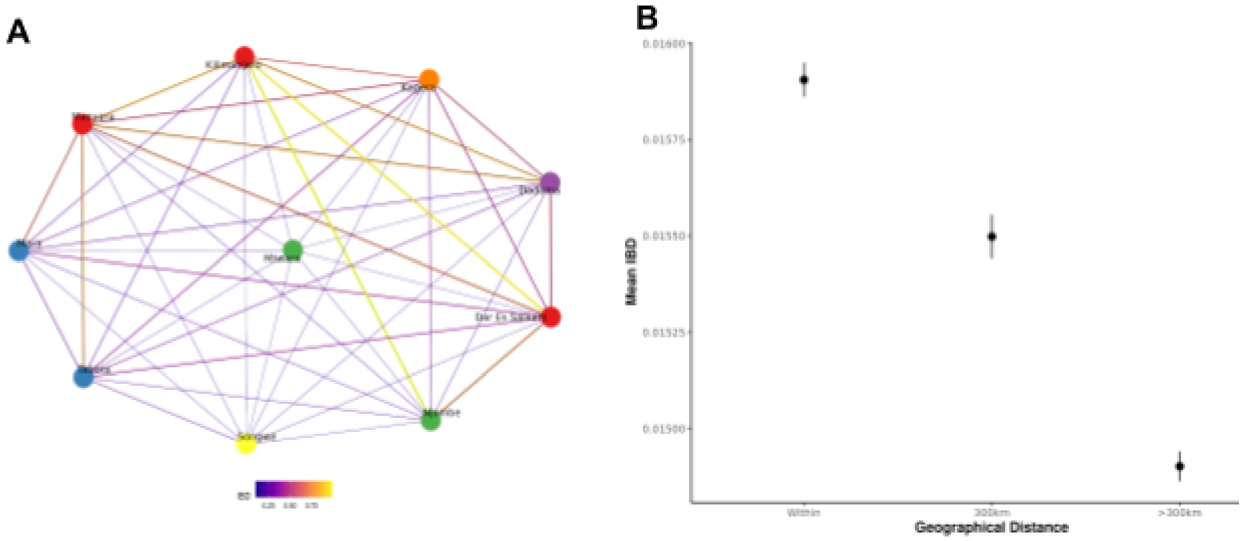

**Figure 4: A)**The effect of spatial distance on Parasite IBD sharing among health facilities and IBD-relatedness decreased as the geographical distance increased. **B)** Parasite population connectivity among 10 regions. Parasites from Kilimanjaro and Manyara exhibit higher connectivity with parasites in other regions

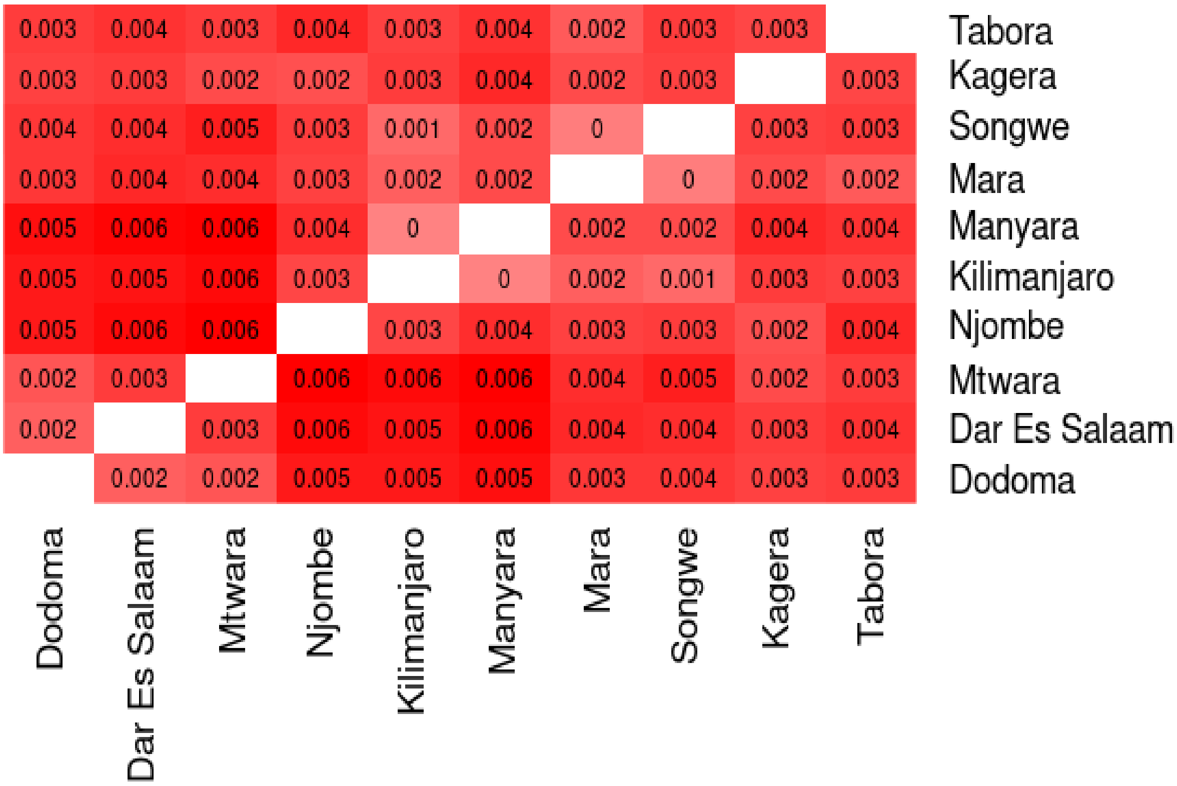


**Figure 5:** The heatmap showing parasite genetic differentiation among regions using pairwise $F_{ST}$ with $F_{ST}$ values ranging from 0 to 0.006 between regions
